# Feasibility and acceptability of daily testing at school as an alternative to self-isolation following close contact with a confirmed case of COVID-19: A qualitative analysis

**DOI:** 10.1101/2021.10.05.21264548

**Authors:** Sarah Denford, Lauren Towler, Behiye Ali, Georgiana Treneman-Evans, Rachael Bloomer, Tim Peto, Bernadette C Young, Lucy Yardley

## Abstract

**Background:** Daily testing using a rapid Lateral Flow Device (LFD) has been suggested as an alternative to self-isolation. A randomised trial comparing daily contact testing (DCT) in schools with self-isolation found that SARS-CoV-2 transmission within school was comparable and low in both groups. However, if this approach is to be adopted widely, it is critical that we understand the perspective of those who will be delivering and receiving DCT. The aim of this qualitative process study embedded in the randomised controlled trial (RCT) was to improve understanding of a range of behavioural factors that could influence implementation.

**Methods:** Interviews were conducted with 63 participants, including staff, students, and parents of students who had been identified as being in close contact with someone with COVID-19. The topic guide explored perceptions of daily testing, understanding of positive and negative test results, and adherence to guidance. Data were analysed using an inductive thematic approach.

**Results:** Results were organised under three main headings: (1) factors influencing daily testing (2) interpretation of test results (3) behaviour during testing period. Participants recognized that daily testing may allow students to remain in school, which was viewed as necessary for both education and social needs. Whilst some felt safer as a result of daily testing, others raised concerns about safety. Participants did not always understand how to interpret and respond to test results, and although participants reported high levels of adherence to the guidance, improved communications were desired.

**Conclusion:** Daily testing may be a feasible and acceptable alternative to self-isolation among close contacts of people who test positive. However, improved communications are needed to ensure that all students and parents have a good understanding of the rationale for testing, what test results mean, how test results should be acted on, and how likely students are to test positive following close contact. Support is needed for students and parents of students who have to self-isolate and for those who have concerns about the safety of daily testing.

## Background

At the time that this study was conducted in the UK, when a case of COVID-19 was identified within a school or college all close contacts of the case were required to self-isolate at home for 10 days. In some situations, the number of close contacts could be quite large. Self-isolation could therefore have considerable negative impacts on the education, psychological health and wellbeing of those affected [1]. It is also possible that preventing students from attending school may not lead to effective isolation, since there is some evidence that suggests that compliance with self-isolation outside the school setting may be as low as 11% in asymptomatic contacts [2].

Daily testing using a rapid Lateral Flow Device (LFD) has been suggested as an alternative to self-isolation. This involves close contacts of a confirmed case being offered the option to take an LFD every day for up to seven days. Tests are taken at school at the start of the school day, and those with a negative test result can remain in school. A person who tests positive with LFD follows national protocols and self-isolates for ten days. This approach has been piloted elsewhere [3] with evidence suggesting that it may be acceptable and feasible [3-5].

Data from a randomised trial of daily contact testing (DCT) in schools carried out during a period of high infection rates in the UK suggests that DCT was not inferior to self-isolation for controlling transmission of SARS-CoV-2 within schools [6]. However, if this approach is to be implemented widely, it is critical that we understand the perspective of those who will be delivering and receiving DCT. The aim of the qualitative process study reported here, which was nested within the RCT, was to improve understanding of a range of behavioural factors, including reasons for participating, response to negative and positive test results, and compliance with self-isolation.

## Methods

### Design

We conducted interviews with students, parents, and staff in a sample of the schools involved in daily testing.

### Sampling and data collection

We asked a key contact in all schools that were involved in the trial of daily testing to invite staff, students, and parents of students who had been identified as being in close contact with a confirmed case of COVID-19 to take part in an interview with the research team. Interested participants were directed to an online form where they provided their contact details, the name of their school, the year group of the student, and whether or not they/their child had taken part in daily testing. We then used a purposive sampling strategy that aimed for diversity in whether or not the participant had taken part in daily testing, school size and location, and year group of the student. We sampled from a range of locations to ensure we sampled participants from schools with high infection rates (including new variants), as infection rates varied considerably between school districts and were typically higher in districts with greater deprivation. Selected participants were contacted by email and provided with an information sheet about the study.

Interviews were conducted remotely (online or by telephone) by a trained qualitative researcher from the University of Bristol (SD, LT, GTE, BA, and RB). Our initial topic guide was designed to explore experiences of the testing process, beliefs about testing, perceptions of positive and negative test results, and impact of testing on behaviour. In order to encourage participants to speak openly about their views and behaviour during the testing period, participants were informed that the interviews would be anonymous and their behaviour would not be disclosed even if they had not always adhered to the guidance. However, participants were reminded that the research team would be obliged to notify authorities if the participant revealed any intended or planned breaches of COVID-19 regulations that could put others in danger.

All participants provided verbal consent/assent prior to taking part in the interview. Parental consent was obtained from parents of all participants under the age of 16 years. Ethical approval was granted by Public Health England’s Research Ethics and Governance Group (ref R&D 434).

### Analysis

Interviews were audio-recorded, transcribed verbatim and anonymised. Data were analysed using an inductive thematic approach [7]. Accordingly, transcripts were read repeatedly by two authors (SD, LT), and detailed notes were made about interesting concepts and ideas. Transcripts were then imported into NVivo software and all text was labelled with an initial set of codes. Team members met regularly to discuss codes and develop a preliminary set of themes. Drawing on existing literature, themes were refined and similar themes grouped together. Charts were then developed for each theme, with the relevant data displayed. The team then explored patterns within and across the full range of participants and groups. Divergent cases were discussed and included in the analysis.

## Results

Sixty-three participants took part in an interview including 24 students, 24 parents and 15 members of staff from 20 schools. Schools were located in Bedfordshire (1); Cheshire (1); Devon (2); Gloucestershire (1); Kent (1); Lancashire (2); Leicestershire (2); Merseyside (1); Northamptonshire (1); Northumberland (1); Oxfordshire (1); Surrey (1); Wiltshire (1); Yorkshire (3); Rutland (1). A total of seven parents, six students and two members of staff did not participate in daily testing.

### Results of the thematic analysis

Data are presented under three main headings: (1) factors influencing acceptability of daily testing (2) test results (3) behaviour during testing period (Table 1).

**Table 1:**
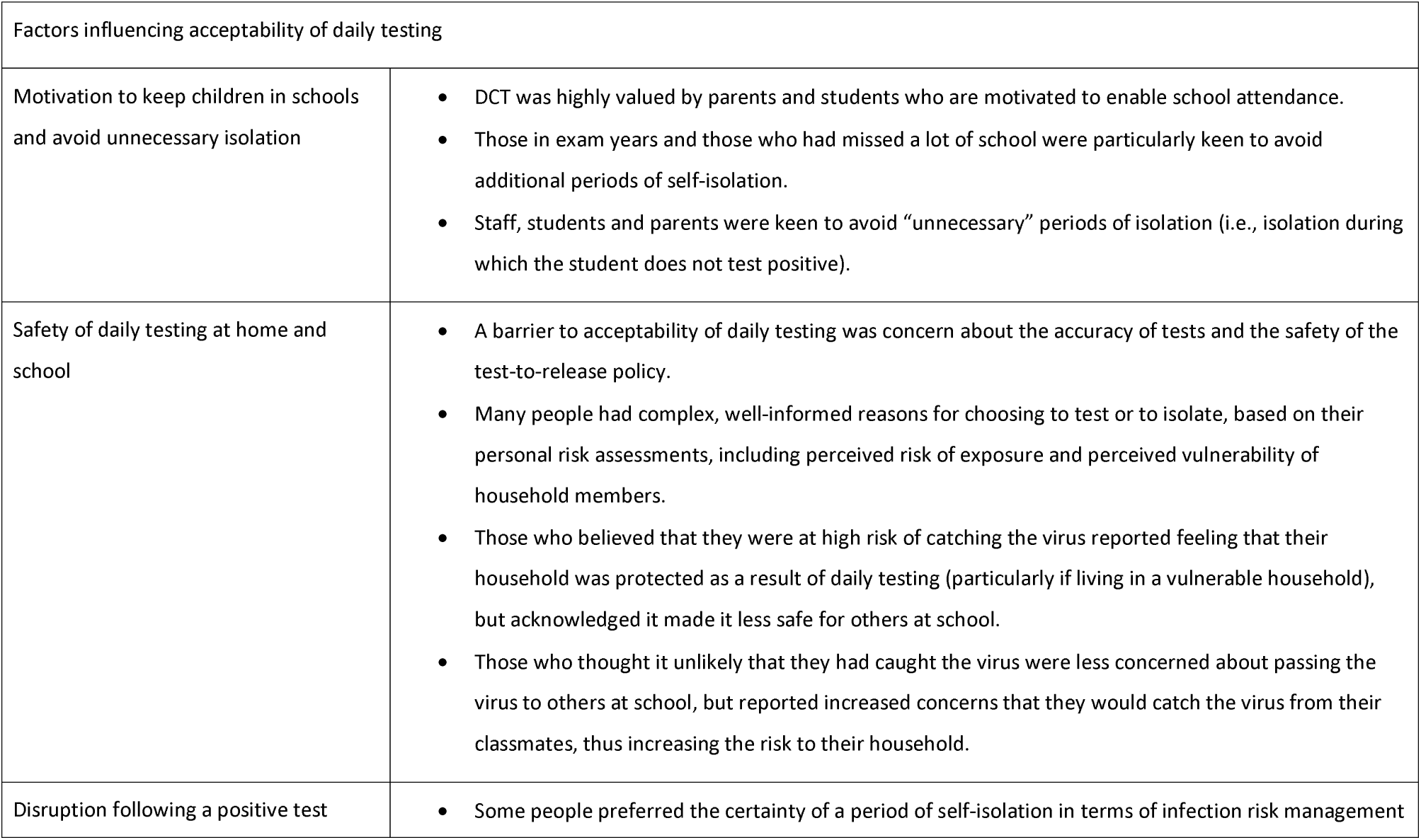
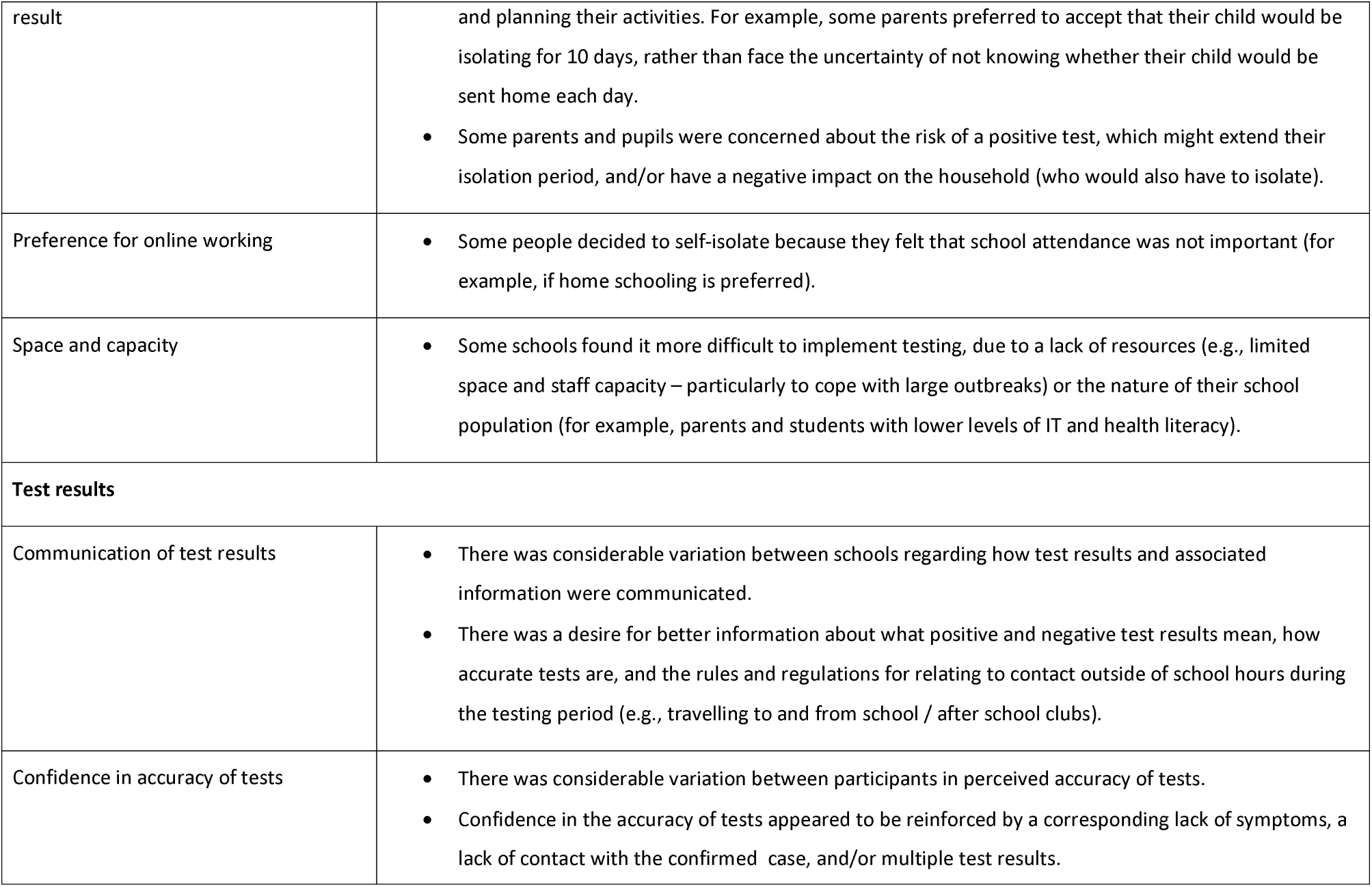
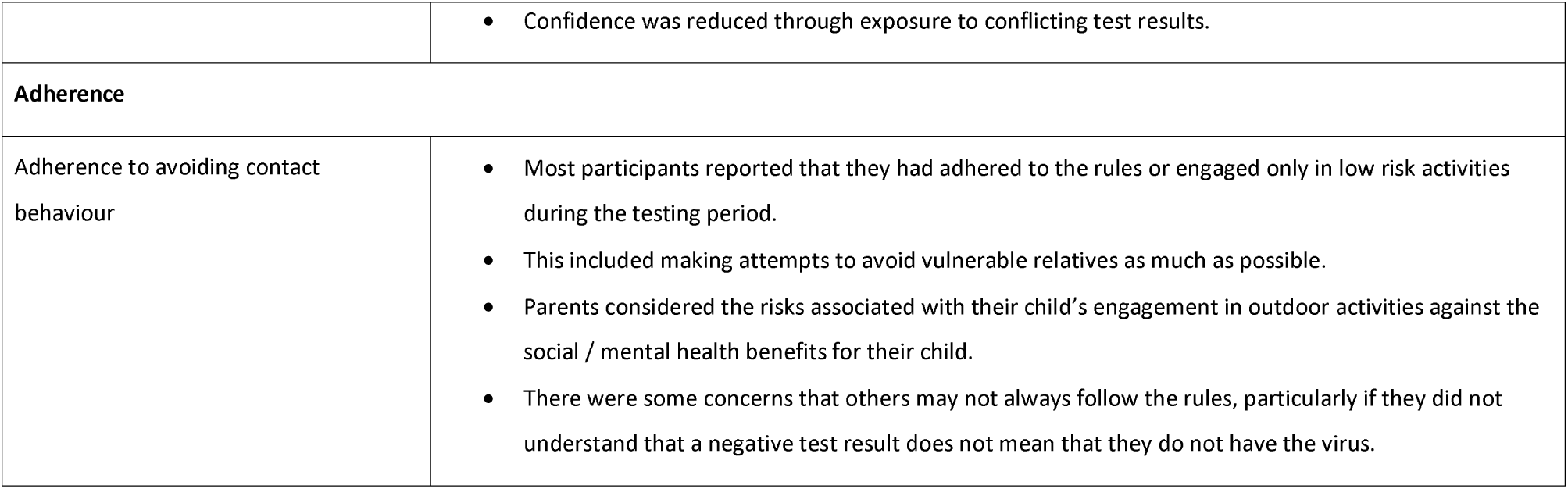
Overview of themes

### Factors influencing acceptability of daily testing

#### Motivation to keep children in school

Involvement in daily testing appeared to be largely driven by the desire to keep children in school in order to fulfil both educational and social needs:

> *“I was quite happy to go into school and socialize. I needed that socializing” (C04, Student, participated in testing)*.

Those in exam years and those who had missed a lot of school over the previous year were particularly keen to stay in school as much as possible:

> *“It was mainly because I didn’t want to get behind in my lessons and in exams because I’ve got exams coming up, so I don’t want to miss out on much learning so I wanted to be at school” (C11, student, participated in testing)*.

Parents and school staff expressed concerns for their children’s mental wellbeing during periods of isolation, and were keen to get their children back to school for the benefit of their mental health:

> *“The emotional and the mental welfare [of isolation] yes, it has because he’s become a little bit more introvert… now he’s gone back to school he’s enjoying it more” (P19, parent, participated in testing)*.
>
> *“I am the one who has to deliver the letters to the students who have to isolate when they are identified as direct and close contact and giving students letters sometime on the third or fourth go that they’ve had to isolate it’s heart breaking when they’re crying you know they don’t want to go home” (S06, Staff, participated in testing)*.

Many students had had multiple periods of isolation in the past, despite never having tested positive, and staff, students and parents were keen to avoid additional, potentially unnecessary time away from school:

> *“I’ve personally had to self-isolate twice. I’ve never tested positive but I’ve had to self-isolate and it’s the worse feeling. I’m an outdoors person so staying indoors for me and doing absolutely nothing is the worst thing” (C06, Student, participated in testing)*.
>
> *“We ended up with about 450-500 students self-isolating in that final month. And it was just like a revolving door with students coming and going so if there’s anyway where we can be part of the pilot where we can offer daily contact testing and hopefully prevent a recurrence of that, that’s the kind of motivation for us in terms of getting on board with this and seeing it as such a high priority” (S01, staff, participated in testing)*.

#### Safety of daily testing

There was considerable variation in participants’ perceptions of the safety of daily testing. One of the biggest concerns among participants was the potentially increased risk of transmission during the testing period. This included concerns about students passing the virus to others, and students being at a greater risk of catching the virus from others during the testing period:

> *“You’re sending them back in where there is a higher risk of catching it because some of those people have been in contact with somebody who’s tested positive” (P14, parent, participated in testing)*.
>
> *“He still was allowed to school on the bus, which defeats the object when it’s a packed bus to get to school” (P3, parent, participated in testing)*.

Concerns about transmission resulted in some participants describing the initiative as unsafe:

> *“I think you’ve obviously picked up the fact that it’s not a programme that I agree with…. I just didn’t feel it was safe” (S12, staff, did not participate in testing)*.

However, whilst some felt the option of testing to release from self-isolation was unsafe, others reported feeling safer as a result of the testing procedures:

> *“My dad has had a stroke and my mum has another health related problem so it’s nice to feel that we’re safe at home and we’re not going to pass on the virus to our parents” (C04, student, participated in testing)*.

Perceptions of safety appeared to depend, to some extent, on whether or not the participant thought it likely that they had caught COVID-19 from the confirmed case. For example, those who thought it possible that they had caught the virus appeared less concerned with catching COVID-19 at school, and felt that participating in testing was a safe way of protecting the household:

> *“[it gave us extra reassurance as a household] yeah, because she was a very close contact with this girl so we knew that at any one point she could be testing positive*” (P18, parent, participated in testing).
>
> *“At the end of the day we thought, well he’s already been in contact with the child so why not keep going to school? And we know we’re safe and secure then, or at least as best we know we’re safe and secure” (P6, parent, participated in testing)*.

However, whilst testing of staff and students who thought they could have the virus was viewed as safer for the household, it was considered to be less safe for others at school:

> *“Obviously, the virus kills people… I felt that by not isolating that I was putting my colleagues at risk because potentially I could have become unwell” (S12, staff, did not participate in testing)*.

This was particularly true for those working with vulnerable individuals:

> *“I was more worried for my colleagues who were older or had other health conditions” (S09, staff, participated in testing)*.
>
> *“I’ve got to think about my work colleagues. In the office that I share I’ve got two of my colleagues that are clinically vulnerable and I’ve also got a colleague who’s pregnant, so I think it would be incredibly selfish if I came to work” (S12, staff, did not participate in testing)*.

Other participants reported that it was highly unlikely that they had caught COVID-19 from the confirmed case, either because they had only seen the person briefly and/or because they had not been very close to them. This group considered themselves likely to be free of COVID-19, and potentially at risk of catching the virus from other close contacts at school. This group considered testing to be less safe for their household, but also noted that there was less risk of them giving the virus to others at school:

> “*If she then stayed in school with all those other children who had been with the person who had tested positive, she would be more likely to then contract it herself… obviously it has knock-on implications for the rest of the family” (P07, parent, participated in testing)*.
>
> *“We don’t think you’ve got it so you weren’t that close a contact, so let’s do the daily [testing]” (P3, parent, participated in testing)*.

Any increase in community cases and/or removal of safety measures in schools could reduce confidence in the safety of testing:

> *“This might be the way that we do have to return to normal life but I do certainly think that the numbers have got to be a lot lower before maybe I would be comfortable… but I think at the moment in time the demographics of our school, the numbers were quite high as well, I just didn’t feel it was safe” (S12, staff, did not participate in testing)*.
>
> *“Now they’re not wearing masks and mixing, yeah [I have concerns about testing]. There are so many holes in the testing regime that you could drive a bus through them… whilst we’ve got the testing regime, I don’t think we have the controls in place that mean that the testing regime would work” (S05, staff, participated in testing)*.

However, the introduction of infection control measures, and / or having had a vaccine against COVID-19 could increase perceptions of safety:

> *“[I was reassured after] seeing how the school had laid out the hall to be tested, how the steps were taken to follow the procedure properly, how it was kept safe” (S09, staff, participated in testing)*.
>
> *“I’m fully vaccinated. I do my swab testing, twice a week, so I feel quite safe” (S11, staff, participated in testing)*.

Despite this, some parents just felt safer with their child at home:

> *“I prefer just to be in me own little bubble, knowing that I’m safe, my children are safe, my husband’s safe – that kind of thing” (P15, parent, did not participate in testing)*..

#### Preference for online / at home working and learning

In contrast to those who had struggled with online learning and were keen to get back to school, other students had enjoyed remote learning and were happy to go back to online learning when they had the opportunity:

> *“So I thought being at home, like I work better at home anyway, so I thought that it would be better at home to revise for my tests coming up” (C13, student, did not engage in testing)*.
>
> *“During lockdown, I enjoyed the live lessons” (C17, student, did not engage in testing)*.

Staff that were able to work from home preferred this option to reduce any unnecessary risk:

> *“I could work from home so, for me the benefits of isolating myself outweighed the need to be in work” (S15, staff, participated in testing)*.

Parents and teachers reported that some students were keen to avoid a week of school:

> “*He is really quite happy about it because he then likes to not have to get up in the morning. He doesn’t have to get on a school bus, and he can do his lessons from home and so, from his point of view, it’s great” (P24)*.
>
> “*A lot of the kids in the group didn’t really want to go to school anyway, so they I think they use it as a bit of an excuse not to have to go to school” (S05, staff, participated in testing)*.

It was thought that age and ability group may influence this:

> *“If I was in Year nine or below, I’ve got to isolate, I would be quite happy… but when it gets to the older years, I’ve got a feeling that I would personally want to do daily testing” (C23, student, participated in testing)*.
>
> *“I just got the impression that [those choosing to isolate] were in a lower end [group]. It wasn’t a top set year group… It was a middle of the road or lower, so a lot of the kids in the group didn’t really want to go to school anyway” (S05, staff, participated in testing)*.

#### Disruption following a positive test result

A number of participants were reluctant to take part due to concerns about receiving a (true or false) positive test result:

> *“I don’t know, but some of my friends have like been falsely positive without any symptoms or anything and I thought it was kind of pointless to keep on doing tests if one of them was going to turn out positive anyway” (C13, student, did not participate in testing)*.
>
> *“I was a bit nervous in case I got a positive test and then I’d have to self-isolate” (C16, student, participated in testing)*.

Some parents preferred the certainty that comes with isolation over the uncertainty of daily testing:

> *“I think my main reason was kind of consistency because obviously they’d have to do the daily testing and then could or couldn’t go in depending on the result of it and it just seemed easier just to go right, we’ll just do that then and then we all know where we are” (P12, parent, did not participate in testing)*.

#### Space and capacity

Conduct of testing appeared to be very much influenced by the amount of (inside and outside) space and staff available, as well as the number of students requiring tests:

> *“We’re very fortunate to have [separate buildings]; it’s completely separate to the main college so we’re able to do all the testing over there. So actually the risk to the bigger school is almost mitigated because we’re able to do that contact testing before they even come onto college site, so you know, we’re very, very fortunate in that. I’m sure many schools aren’t*” (S02, staff, participated in testing).
>
> *“So we’re very small… I wouldn’t want to do it in a mainstream secondary to be honest having worked in mainstream secondaries, the thought of queues snaking around the building in the freezing cold and the rain waiting to come into a particular area yeah I could see that would be much more difficult to manage” (S08, staff, participated in testing)*.

Depending on the resources that were available, schools adopted different approaches to testing, however, staff resources were a restrictive factor:

> *“We haven’t got spare staff to be doing any of these jobs” (S03, staff, participated in testing)*.

Regardless of the size of the school, it was not considered possible to test a large number of close contacts when demand outweighed capacity:

> *“We had this one last week… we just said we’re really sorry but we have got to capacity, I think we’ve got 150 students that the girls are having to get through and I think that was our limit… and we can’t drag anybody from anywhere else cos we haven’t got anybody otherwise it’s impacting on teaching and learning” (S06, staff, participated in testing)*.

### Test results

#### Communication of test results

Although study procedures required the results to be communicated (with a script provided), there was considerable variation between schools in how information about tests and test results were disseminated. Due to workload and resources, many schools opted to only inform students and parents of positive test results. This meant that students and parents did not receive any information about how to interpret tests and the associated rules and regulations at the time of testing:

> *“What we said to our parents is we’d only communicate if it was a positive test, so the negative tests we haven’t communicated” (S08, staff, participated in testing)*.
>
> *“No [information about test results are shared at the time of testing], because at that point they’ve already completed the consent form for us, so they already know what the tests are about, etc” (S4, staff, participated in testing)*.

This led to variations between school staff regarding how confident they were that students and parents had received all the information needed to follow the rules:

> *“Well we’ve been quite explicit about explaining that the reason why… and like I say the vast majority of people are abiding by it. So yeah I think everything’s quite easy to understand really” (S6, staff, participated in testing)*.
>
> *“We didn’t really communicate apart from to tell the kids that, ‘You’re not positive today. You can attend school etc*., *but you have to self-isolate when you get home’. I get the impression that most of it was lost somewhat on the children” (S05, staff, participated in testing)*.

Parents and students reported that confirmation of test results would have been useful:

> *“You wouldn’t have been able to go and do even just a bit of shopping because you didn’t know if you had to wait around to pick her up” (P2, parent, participated in testing)*.
>
> *“I just thought I’d like to be told if we were positive or negative because when people say ‘Have you tested negative?’ and I say ‘Well I’m not really sure because I’m allowed in, but I haven’t been told actually if I am positive or negative*.*’ You have to just assume” (C1, student, participated in testing)*.

Furthermore, there was evidence that students and parents did not always understand daily testing rules and requirements:

> *“We were quite cross. That was not clear… it wasn’t clear whether actually he could do his extra curricula activities. I actually emailed the school and had to ask them because that was never clear. I think, I don’t know whether a few parents got confused about that because they were saying I’ve seen kids out. [He] couldn’t walk to school with his friends. He had to walk on his own because his friends that he walked with was in a different form group but [he] told me that. That didn’t come from the school. He told me that he wasn’t allowed to walk with friends” (P21, parent, participated in testing)*.

There was evidence that some parents, staff, and students were confused about the logic underpinning the rules:

> *“I didn’t understand that if I tested negative that day, then I was allowed to go to school, why was I not allowed to go to everything else?” (C19, student, participated in testing)*.
>
> *“It makes no sense, because it’s shoulder to shoulder [at school], but they are not allowed to go and see each other and stand two metres away outside [after school]… I can see why they are kind of not [adhering to the rules]” (S09, staff, participated in testing)*.

#### Confidence in accuracy of tests

Whilst levels of confidence in the accuracy of testing varied, participants typically acknowledged that tests were not infallible:

> *“[I was] quite confident but a little bit not confident” (C16, student, participated in testing)*.
>
> *“I think the tests are what, 50% accurate or something, 60. I don’t know what the number is, but I know it’s not 100%” (S05, staff, participated in testing)*.

Despite general agreement that tests were not always accurate, those participating in daily testing were usually confident that the tests were “good enough” to facilitate a safe return to school:

> *“I know there’s been a lot of talk about the lateral flow tests maybe not being a great indicator but I still think it’s you know it’s a better alternative than her not being there (P01, parent, participated in testing)*.
>
> *“We’ve got very few staff who are not wanting to do the test because they feel that so long as it’s more than 50% accurate, just to swab your nose and your throat and to get some kind of result that could be more than 50% accurate is better than not testing at all. …So absolutely everybody who does that test understands that it’s not 100% accurate. I’ve said it was 70-odd% accurate, the one we were previously using, and everybody that I’ve spoken to has said yeah, that’s better than doing nothing” (S03, staff, participated in testing)*.

Some participants were willing to trust the tests – simply because the process was being implemented in, and supported by the school or trial management committee:

> *“Somebody somewhere must be [confident that the tests were accurate] to have put the trial in place, so I guess I kind of bow to their thought process” (P05, parent, participated in testing)*.

Confidence in the accuracy of tests appeared to be reinforced by a corresponding lack of symptoms, a lack of contact with the confirmed case, and/or multiple test results:

> *“I was pretty confident because I had barely been near the person. In my seating plan the tables are very far apart so I wasn’t actually that close to her and I’ve never actually walked passed her or spoken to her or anything, so I was pretty confident that they were reliable and true” (C11, student, participated in testing)*.
>
> *“I was pretty confident because obviously they were the same each day, so if I was positive, it would have shown up on one of the days at least probably” (C08, student, participated in testing)*.

Repeated exposure to concordant test results enhanced confidence:

> *“Every time we’ve had a positive, we’ve ended up with a complimentary PCR positive also. So I’m fairly confident they’re accurate. They have been for us, anyway (S4, staff, participated in testing)*.

However, exposure to discordant test results appeared to weaken confidence:

> “*I’m not 100% sure of the validity of the test because for example my brother got tested negative and then one day later he got COVID-19. Since then I’ve been quite sceptical and not 100% sure about what it is” (C04, student, participated in testing)*.

In such cases, students employed strategies such as multiple consecutive tests to increase their confidence in the results:

> *“I had already taken three the night before… um, because the person that tested positive said that she had done four and two came back positive and two came back negative, so I just did three to make sure” (C19, student, participated in daily testing)*.

#### Adherence to avoiding contact

Most students and parents reported either that they had adhered to the rules, or had engaged in low to no contact activities (e.g., taking the dog for a walk, seeing friends they were at school with):

> *“I took my dog on dog walks but we just went around my estate, we didn’t really see anyone” (C11, student, participated in testing)*.
>
> *“The only time that he was out of the house outside of going to school was we went for a walk one day somewhere where there wasn’t anybody else, so there was no risk of passing on to anybody else and he’d also tested negative the day before, so we knew that the risks were very minimal. And I know in theory according to the rules we shouldn’t even have done that, but we did just do that one walk, because we all needed it” (P05, parent, participated in testing)*.

Behaviour within the home appeared to be very much dependent on the risk status of the household, with those living in high-risk households wanting to avoid contact as much as possible during the testing period:

> *“I just tried to keep my distance a bit more because I’ve got my 80 year old grandma living with us at the moment so obviously trying to keep myself away as much as possible was a priority really because obviously I don t want her to get it if I had it. I tried to keep my distance a bit more” (C01, student, participated in testing)*.

Parents reported actively weighing up the risks associated with their child’s engagement in outdoor activities against the need for testing to have some benefit for the children:

> *“Having read it at the time we just thought that he goes to school, he’s having a daily test, so that means life carries on as normal… Then it was a couple of days later when we read it again and it’s life doesn’t carry on as normal but it gives you the ability to go to school but you shouldn’t be doing anything else. That didn’t really sit comfortably with us … It felt very much, other than school, where’s the gain, really? When things are just starting to open up, he’s been a week into playing football, he plays football for a team that had just started training again, so to be fair, he went to football… he’s having [a test] every day, that should be, in my mind and in my husband’s mind. Daily test, that gives you the freedom to live normally… I suppose we just took the view that we can’t afford for ((name)) to miss anymore of his socialisation in his life” (P8, parent, participated in testing)*.

Indeed, parents appeared to weigh up additional dis/advantages of ‘breaking the rules’ in terms of the effect it would have on their child’s mental health, even though it may not be permitted:

> *“She was a bit frustrated saying, ‘Everyone’s going out,’ and we had a talk about what the situation might be if her friend came out, and if it was friends she’s at school with and they were going to be outside in the open air, and friends were okay with it, and there weren’t too many of them, and they’re people that she’d been mixing with at school. I said, ‘We can discuss it if that is the case,’ thinking maybe there’d be some pressure, that she’ll want to join them and maybe that wouldn’t be stretching the rules too much” (P11, parent, participated in testing)*.

One student described receiving pressure from her employer to attend work, possibly resulting from a lack of awareness and understanding of multiple testing initiatives:

> *“I had to go to work because the student didn’t give our names to Track and Trace, so I still had to go to work over the weekend and one day during the week*… *[my boss] said as long as you didn’t have a message from Track and Trace you’re fine*” *(C19, student, participated in testing)*.

Whilst participants largely described adhering to the rules themselves, there were some concerns that others may not always do so:

> *“I think that’s where the real danger will be, is that people will think that daily contact, you know, that daily test, gives me 24-hours for want of a better word, protection. And they think a) it means I’m virus free and b) I don’t need to worry about any kind of, you know, wider behaviour outside of school” (S01, staff, participated in testing)*.

## Discussion

### Summary of findings

To our knowledge this is the first study to qualitatively explore perceptions of daily testing in schools as an alternative to self-isolation following close contact with confirmed cases of COVID-19. Our work reveals that use of DCT may be supported by those who are motivated to enable school attendance and avoid self-isolation, and who perceive the risks of testing to be low for them and their contacts. However, we also identified a number of situations in which self-isolation may be preferred. In particular, staff and students who favour home working/studying, were concerned about disruption following a positive test result, or were concerned about transmission of the virus during the testing period were more likely to opt to self-isolate.

Previous research suggests that staff, students and parents are concerned about perceived risks of missed learning [8], and that these concerns may outweigh concerns about transmission of the virus. In the current study, many participants were keen to avoid periods of isolation and remain at school. In particular, students in critical (exam) years, those who value school attendance, and those who had experienced previous periods of self-isolation appeared most accepting of DCT as an approach to reduce periods of school absence. However, some participants were not convinced that the need to keep students in school did outweigh the risks of transmission in schools, and for these individuals additional information and reassurance about the safety of testing may be needed. Furthermore, some students described substantial benefits of remote learning, appreciating the opportunity to work at their own pace in an environment free of distractions.

Whilst previous work identified support for COVID-19 testing in schools [8] it was not clear whether daily testing following close contact with the virus would be equally supported. In the current study, perceptions of the safety of DCT varied considerably between participants, and was often based on a complex and sophisticated evaluation of their perceived risk of exposure and the perceived vulnerability of those around them. Some students thought that the risk that they had already caught COVID-19 was high due to close contact with the confirmed case and so did not think attending school would increase their risk, but they wanted the reassurance of a daily test result to confirm that they were not transmitting the virus to members of their household. However, those considering themselves likely to have already caught COVID-19 could be motivated to avoid schools (and self-isolate) if in close contact with vulnerable staff and students within the school setting. Likewise, some students decided to self-isolate because they thought they were unlikely to have caught COVID-19 from the confirmed case, and considered themselves/their households to be at greater risk if they attended school and were in close contact with others who may have COVID-19. Conversely, other students who considered themselves unlikely to have contracted the virus could actually demonstrate an increased willingness to attend school, considering the perceived lack of contact with the person who tested positive, as they considered themselves unlikely to be of risk to others.

In line with previous research [5], some parents and pupils were concerned about the risk of a positive test, which could extend their isolation period, and/or have a negative impact on the household (who would also have to isolate). In some cases, accepting that the child would be isolating for 10 days was preferable to facing the uncertainty of not knowing whether their child would be sent home each day. Due to limited resources, schools were not always able to communicate test results with parents, and this often added to the uncertainty and prevented planning. Improved communications between schools and parents may reduce the uncertainty to some degree. Furthermore, evidence suggests that rates of COVID-19 in school-based contacts are very low [6]. Communication of such findings may alleviate concerns about having to isolate following positive test results. There is also a continued need for improved support for those who do need to self-isolate following positive test results.

Whilst nearly all participants accepted that tests were not conclusive evidence of COVID-19 status, a range of estimates were given regarding their accuracy to predict presence or absence of the virus. Participants were more likely to accept a negative test result as accurate if they had not been in close contact with the confirmed case, had no symptoms, and had taken multiple tests (with the same result). Confidence was weakened through exposure to dis-concordant test results. In order to increase confidence in testing, some participants reported behaviours such as taking multiple LFD tests or a PCR test. Previous research has also highlighted confusion over which tests should be used and when [9]. Provision of accessible information regarding the best way to use tests and how to interpret and respond to uncertain or conflicting test results could help increase understanding, confidence and appropriate use of tests.

Many parents and students had a good understanding of the risks and limitations of daily testing, and the implications for managing the residual risk to others. Our research is consistent with previous qualitative research that also suggested that testing did not increase high risk behaviour [5]. Indeed, in the current study, the majority of participants reported adhering to the guidance or only engaging in low-risk activities, with efforts being made to avoid contact with vulnerable individuals. However, research conducted by the Office for National Statistics found that parents were concerned about the effects of isolation on their child’s mental wellbeing [10], and this could lead to non-adherent behaviour for the sake of the child’s welfare [10]. Similarly, within the current study, breaches of regulations appeared to be the result of an assessment of the dis/advantages of ‘breaking the rules’ in terms of the effect it would have on their child’s mental health, whilst simultaneously attempting to minimise risk as much as possible.

Much research has highlighted the detrimental impact of the introduction of infection control measures without sufficient explanation and justification [11]. Within the current study there was evidence of confusion about why behaviours were permitted in some settings and not others, and how to interpret apparently contradictory test results. Without further clarity and explanation, these perceived mixed messages may weaken adherence overtime [11, 12]. Accessible information about what positive and negative test results mean, how accurate tests are, and the rules and regulations for relating to contact outside of school hours during the testing period may be critical.

### Limitations of the study

The main limitation of this study is that, despite purposive diversity sampling, it is likely that people with more negative views of and less engagement with daily testing may have been under-represented in our sample as they would be less likely to engage with research. Our findings must be interpreted with this in mind.

### Implications for policy and practice

Many students, staff and parents are motivated and competent to implement daily testing appropriately to enable school attendance. However, the acceptability, feasibility and effectiveness of implementing a policy of daily testing may vary between households and schools, and may also vary depending on the context of local infection levels (since these affect perceptions of the risks involved). Better and more accessible communications are needed to ensure that all students and parents have a good understanding of the rationale for testing, what test results mean (including their level of accuracy and limitations), how test results should be acted on, and how likely students are to become infected following close contact. Particular attention is needed to improve acceptability and feasibility of testing in schools and households with lower levels of IT literacy and health literacy.

The option of daily testing of contacts may be beneficial for those who require reassurance to reduce anxiety about transmission from school contacts to vulnerable household members. However, concern about a positive test resulting in a longer self-isolation period or the need for other household members to self-isolate poses a barrier to the acceptability of testing, especially in households with less resources for self-isolation.

## Data Availability

The datasets used and/or analysed during the current study are available from the corresponding author on reasonable request.

## Conclusions

This study suggests that DCT may be a feasible and acceptable alternative to self-isolation among close contacts of COVID-19 cases. Participants were motivated to keep children in school but raised concerns about the safety of daily testing in some circumstances. Improved communications are needed to ensure that all students and parents have a good understanding of the rationale for testing, what test results mean, how test results should be acted on, and how likely students are to be a case following close contact. Support is needed for students and parents of students who have to self-isolate and for those who have concerns about the safety of DCT for their household.

## Declarations

### Ethics approval and consent to participate

Ethical approval for this study was granted by Public Health England’s Research Ethics and Governance Group (ref R&D 434).

### Consent for publication

All participants provided verbal consent for data to be included in publications

## Acknowledgements

LY is an NIHR Senior Investigator and her research programme is partly supported by NIHR Applied Research Collaboration (ARC)-West, NIHR Health Protection Research Unit (HPRU) in Behavioural Science and Evaluation, and the NIHR Southampton Biomedical Research Centre (BRC).

SD is supported by the NIHR Health Protection Research Unit (HPRU) in Behavioural Science and Evaluation at the University of Bristol in partnership with Public Health England.

BCY, TEAP, SD and LY received grants from the Department of Health and Social Care to fund this work.

## References

1. H.M Government. Academic Year 2020/21: Schools, Pupils and their characteristics 2021: https://explore-education-statistics.service.gov.uk/find-statistics/school-pupils-and-their-characteristics. [Accessed July 2021]

2. Smith, LE, Potts HW, Amlot R, Fear NT, Michie S, Rubin GJ. Adherence to the test, trace and isolate system in the UK: results from 37 nationally representative surveys (the COVID-19 Rapid Survey of Adherence to Interventions and Responses [CORSAIR] study). BMJ, 2021. 372;n608.

3. Love N, Ready D, Turner C, Yardley L, Rubin J, Hopkins S, Oliver S. The acceptability of testing contacts of confirmed COVID-19 cases using serial, self-administered lateral flow devices as an alternative to self-isolation. MedRxiv [preprint] (2021). Available at The acceptability of testing contacts of confirmed COVID-19 cases using serial, self-administered lateral flow devices as an alternative to self-isolation | medRxiv

4. Martin AF, Denford S, Love N, Ready D, Oliver I, Amlot R, Rubin J, Yardley J. Engagement with daily testing instead of self-isolating in contacts of confirmed cases of SARS-CoV-2. BMC Public Health, 2021. 21(1067): doi.org/10.1186/s12889-021-11135-7

5. Denford S, Martin F, Love N, Ready D, Oliver I, Amlot R, Yardley J, Rubin J., Engagement with daily testing instead of self-isolating in contacts of confirmed cases of SARS-CoV-2: A qualitative analysis. Frontiers in Public Health, 2021. 9:1109 https://doi.org/10.3389/fpubh.2021.714041

6. Daily testing for contacts of individuals with SARS-CoV-2 infection and attendance and SARS-CoV-2 transmission in English secondary schools and colleges: an open-label, cluster-randomised trial. Lancet, 2021 Sep 14:S0140-6736(21)01908-5. doi: 10.1016/S0140-6736(21)01908-5

7. Braun, V. and V. Clark, Reflecting on reflexive thematic analysis. Qualitative Research in Sport, Exercise and Health, 2019. 11(4):p. 589–597.

8. Lorenc, A., Kestern, JM., Kidger, J., Langford, R., Horwood, J. Reducing COVID-19 risk in schools: a qualitative examinaiton of secondary school staff and family views and concerns in the South West of England. BMJ Paediatric Open, 2021. 5(1): p. e0000987.

9. Smith, L., Potts, HWW., Amlot, R., Fear, NT., Michie, S., Rubin, GJ. Do members of the public think they should use lateral flow tests or PCR tests when they have COVID-19-like symptoms? The COVID-19 Rapid Survey of Adherence to Interventions and Responses [CORSAIR] study. Public Health, 2021. 198:260–262 https://doi.org/10.1016/j.puhe.2021.07.023

10. Office of National Statistics. In their own words: How different people respond to coronovirus guidance. 2021. [Accessed July 2021]

11. Burton, A., McKinlay, A., Dawes, J., Roberts, A., Fynn, T., May, T., Fancourt, D. Understanding barriers and facilitators to compliance with UK social distancing guidelines during the COVID-19 pandemic: A qualitative interview study. PsyArXiv preprints, 2021.

12. Williams S, Armitage C, Tampe T, Dienes K. Public perceptions of non-adherence to COVID-19 measures by self and others in the United Kingdom. MedRxiv. 2020. DOI: 10.1101/2020.11.17.20233486.

